# Institutional Distrust as the Most Frequently Reported Barrier to Health Insurance Enrollment in Post-Conflict Liberia: A Mixed-Methods Study Extending the Andersen Behavioral Model with Trust as a Fourth Enabling Resource

**DOI:** 10.64898/2026.11.22.26368698

**Authors:** Olabayo Kehinde Omisore, Benedict Wollor, Vetty Agala, Daprim Ogaji

## Abstract

Health insurance enrollment in post-conflict Liberia remains below 15% despite a decade of reform, and the widely used Andersen Behavioral Model gives institutional trust no explicit place among the determinants of service use. We examined institutional distrust as a theoretically distinct barrier to enrollment using a concurrent mixed-methods design in four counties (Montserrado, Margibi, Nimba and Grand Gedeh). A cross-sectional survey of 542 adults (271 insured, 271 uninsured) ranked self-reported primary barriers and modeled enrollment with multivariable logistic regression; 16 key informant interviews and four focus group discussions, one per county, were analyzed thematically, combining deductive orientation to Andersen’s framework with inductive derivation of trust subtypes, and integrated with survey findings through joint display. Among uninsured respondents, institutional distrust was the most frequently reported primary barrier (57/271, 21.0%), narrowly ahead of premium cost (49, 18.1%) and inaccessibility of services (47, 17.3%), and the barrier distribution did not differ significantly by county. Four analytic subtypes were developed (fund-stewardship, benefit-delivery, provider-quality and generalized-state distrust), although direct evidence for provider-quality distrust overlapped with wider state-health-sector distrust. In multivariable analysis, socioeconomic status was the strongest structural predictor of enrollment: formal employment (adjusted odds ratio [aOR] 8.78, 95% CI 4.90 to 15.76), tertiary education (aOR 8.17, 4.12 to 16.20) and secondary education (aOR 7.36, 3.98 to 13.63) each independently raised the odds of enrollment, whereas informal employment (aOR 1.35, 0.81 to 2.24), sex and county were not significant. Socioeconomic status thus structures who enrolls, while institutional distrust is the barrier the uninsured most often name, a pattern the qualitative data corroborate. We propose extending the Andersen model to treat institutional trust as a fourth enabling resource alongside financial, organizational and social-support resources. In fragile states, trust-building should be treated as health-system infrastructure that complements, rather than replaces, financial-access measures.

**KEY MESSAGES:**

- About one in five uninsured adults in four Liberian counties named distrust of the insurance scheme as their main reason for not enrolling, slightly more than named premium cost, and the overall pattern of barriers did not differ significantly between urban, peri-urban, conflict-affected and remote counties.
- Distrust is not a single attitude. Participants distinguished doubts about whether premiums are safeguarded, whether promised services are delivered, whether care under insurance is of acceptable quality, and whether the state acts in good faith; each points to a different policy lever.
- Education and formal employment remain the strongest predictors of who holds insurance, so trust-building must accompany, not substitute for, measures that lower financial and access barriers for informal-sector households.
- Enrollment strategies in fragile and post-conflict states should budget for trust-building as health-system infrastructure: ring-fenced and audited funds, published benefit guarantees, responsive grievance redress, quality monitoring and inclusive oversight, and studies using the Andersen model should measure institutional trust as an enabling resource.

**Reflexivity statement:** The author team comprises public health and health systems researchers based at Nigerian institutions and a Liberia-based co-author: the first author is a doctoral researcher at the University of Port Harcourt, one co-author is based at a medical center in Liberia and facilitated field access, and the other co-authors bring experience from a state contributory health insurance program (RIVCHPP) and a university center of excellence in public health research. With most of the team working from Nigerian institutional bases, the team was attentive to the risk of importing assumptions from the Nigerian insurance context. Experience within the team of insurance program administration may have sensitized the analysis to scheme design and stewardship concerns. To examine how these positions shaped interpretation, the lead analyst kept reflexive memos throughout coding, community leaders were consulted on instrument wording, and preliminary findings were presented back to community focus groups in each study county before manuscript preparation.

**Ethical approval:** Ethical approval was granted by the Research Ethics Committee of the University of Port Harcourt (December 2024; reference UPH/CEREMAD/REC/MM105/030). A letter of local institutional support for data collection in Liberia was obtained (May 2026). Written informed consent was obtained from all participants. Funding, acknowledgements, conflicts of interest and author contributions are given in the Declarations section.

## INTRODUCTION

Universal health coverage (UHC) commits states to ensure that all people receive the health services they need without suffering financial hardship (World Health Organization 2023). In sub-Saharan Africa, social health insurance and community-based schemes have become the principal mechanisms for advancing this commitment, and countries such as Ghana and Rwanda have shown that meaningful coverage is achievable even at modest income levels (Lu et al. 2012; Wang et al. 2017). Liberia sits at the harder end of this spectrum. Two consecutive civil wars (1989 to 1996 and 1999 to 2003), followed by the 2014 to 2016 Ebola epidemic and the COVID-19 pandemic, have left the health system rebuilding from a near-zero baseline, and enrollment in the country’s nascent social health insurance scheme remains below 15% of the population (Lee et al. 2011; Kentoffio et al. 2016; Ministry of Health, Republic of Liberia 2021).

Persistently low enrollment is conventionally explained through familiar levers: low income, limited information, geographic barriers to access and inadequate service quality at participating facilities (Carrin et al. 2008; Kutzin 2013). These factors map cleanly onto the Andersen Behavioral Model of Health Services Use, which organizes the determinants of care-seeking into predisposing characteristics (demographics, social structure and beliefs), enabling resources (financial, organizational and social-support resources) and need (perceived and evaluated illness) (Andersen 1995; Andersen and Davidson 2014). The model has proven durable across more than four decades of refinement and remains the most widely applied theoretical framework in health services research (Babitsch et al. 2012).

In post-conflict and fragile-state contexts, however, the standard Andersen variables explain less of the variation in enrollment than the model would predict (Newbrander et al. 2011; Witter et al. 2015). Empirical work from Sierra Leone, the Democratic Republic of Congo and other conflict-affected settings repeatedly surfaces a category of barrier that the model does not name explicitly: a generalized and durable distrust of state institutions, of fund custodians and of the health system itself (Pieterse and Lodge 2015; Vinck et al. 2019). Where such distrust is high, even well-designed schemes with subsidized premiums and nearby facilities can fail to attract enrollment (Ozawa and Sripad 2013); where institutional credibility is being rebuilt, enrollment can rise rapidly even amid resource constraints, as the expansion of Rwanda’s community-based health insurance after 1994 illustrates (Lu et al. 2012).

Trust is not absent from health services research. A substantial literature has examined interpersonal trust between patients and individual providers (Hall et al. 2001), trust in vaccination programs (Larson et al. 2018) and trust as a determinant of treatment adherence and service use (Whetten et al. 2006). What remains underdeveloped is the treatment of institutional trust, as distinct from interpersonal trust, as a structural enabling resource on a par with money, organizational capacity and social networks. In the Andersen framework as conventionally operationalized, trust appears, if at all, as a predisposing belief: an attitudinal variable measured by one or two Likert items and modeled as a covariate rather than as a category in its own right (Gilson 2003).

This paper argues that, in post-conflict settings, this treatment is inadequate. Using mixed-methods data from four Liberian counties, we show that institutional distrust is the most frequently reported barrier to enrollment among the uninsured, narrowly ahead of cost; that it is conceptually and empirically distinguishable from cost and access; that it has identifiable subtypes with distinct policy implications; and that it is rooted in experience of institutional behavior rather than in fixed individual attitudes. We also show that distrust operates alongside, not instead of, a steep socioeconomic gradient in enrollment. On this basis we propose a formal extension of the Andersen Behavioral Model that incorporates institutional trust as a fourth enabling resource, alongside the existing financial, organizational and social-support categories.

The paper makes three contributions. First, it provides an empirical mapping of distrust subtypes in a post-conflict African health system, grounded in primary survey and qualitative data from four contrasting counties. Second, it offers a theoretical extension to the most widely used framework in health services research, with a reasoned case for treating trust as an enabling rather than a predisposing factor. Third, it sets out a practical agenda for trust-sensitive enrollment policy in fragile states that distinguishes the interventions suited to each distrust subtype. The study addressed three questions: (1) how frequently do uninsured adults in Liberia name institutional distrust, relative to cost, access and information, as their primary barrier to enrollment; (2) which socio-demographic characteristics independently predict enrollment; and (3) what forms does institutional distrust take, and how can it be located within the Andersen model?

### Conceptual framework

#### The Andersen Behavioral Model: structure and limits

Andersen’s Behavioral Model, first articulated in 1968 and substantially revised in later iterations, posits that an individual’s use of health services is shaped by three sets of factors (Andersen 1995). Predisposing characteristics include demographic features (age, sex), social structure (education, occupation, ethnicity) and health beliefs (attitudes, values and knowledge about health and health services). Enabling resources are the means by which an individual can act on a need: personal and family resources (income, insurance status), community resources (provider availability, regional distribution) and, in later revisions, social support. Need is differentiated into perceived need (the individual’s own assessment) and evaluated need (professional judgment of clinical requirement).

In its mature form, the model accommodates feedback loops: use influences outcomes such as perceived health and satisfaction, which in turn feed back into predisposing characteristics and perceived need (Andersen and Davidson 2014). It has been applied to almost every category of health service and in almost every type of health system, from emergency department use in the United States to outpatient service use in urban China (Babitsch et al. 2012; Bao et al. 2017). Its empirical durability is unusual in the social sciences.

The model’s limits become apparent, however, in settings where the institutional preconditions for service use are themselves contested. Andersen’s framework implicitly assumes a functioning and broadly trusted institutional substrate: that fund custodians will honor claims, that providers will deliver the services for which they are reimbursed and that the state will not redirect pooled premiums to other purposes. Where this substrate is intact, distrust can reasonably be treated as one predisposing belief among many. Where it is not, distrust ceases to be merely a belief about the system and becomes a reasoned assessment of the system’s past behavior: in effect, a missing enabling resource that blocks action regardless of income, information or proximity (Newbrander et al. 2011; Vinck et al. 2019).

#### Why institutional trust is enabling, not predisposing

Treating institutional trust as a predisposing belief, the conventional approach, raises three problems in fragile-state contexts. First, it conflates trust with knowledge or attitude, whereas in post-conflict settings distrust is frequently the product of direct experience of misappropriated funds, denied claims or providers refusing to accept insurance cards (Pieterse and Lodge 2015). It is therefore not a stable trait but a contingent assessment that institutional behavior can raise or lower. Second, it implies that distrust can be remedied through information campaigns or attitudinal change, whereas in practice it responds chiefly to demonstrated institutional reliability over time (Vinck et al. 2019). Third, it separates trust analytically from the resources it most resembles: where the rule of law is partial, trust functions much as money or transport does, as a precondition that must be present for service use to occur.

These characteristics (contingency on institutional behavior rather than individual attributes, responsiveness to structural rather than attitudinal intervention, and functioning as a precondition for use) are the defining features of an enabling resource in the Andersen taxonomy. Treating institutional trust accordingly is therefore not a departure from the model but a clarification of where, within its existing structure, trust most coherently sits.

#### Proposed extension: trust as a fourth enabling resource

We propose extending the enabling-resources category to comprise four parallel subcategories: (a) financial resources (income, insurance and ability to pay); (b) organizational resources (provider availability, a regular source of care and geographic access); (c) social-support resources (family, kin and community networks that facilitate use); and (d) institutional-trust resources (a credible expectation that fund custodians, providers and the state will deliver what enrollment promises). The first three are present in Andersen’s current formulation; the fourth is the proposed addition.

In this formulation, institutional trust has identifiable subtypes that can operate independently and carry distinct policy implications. Drawing on our qualitative data and the fragile-state literature, we distinguish four: (i) fund-stewardship trust, that pooled premiums will be safeguarded and not diverted; (ii) benefit-delivery trust, that promised services will be available when claimed; (iii) provider-quality trust, that participating facilities will deliver care of acceptable quality under insurance; and (iv) generalized-state trust, the diffuse expectation that state institutions act in good faith, which in Liberia is traceable to civil-war-era institutional collapse and uneven post-conflict recovery.

The extension is theoretically conservative: it displaces no existing category, leaves the model’s underlying logic unchanged and remains testable with the model’s standard empirical apparatus. It is nonetheless consequential. It implies that enrollment policy in fragile states should include trust-building as a programmatic line item rather than a soft adjunct, and that measurement of enabling resources should include trust subtypes alongside income and proximity. The remainder of the paper presents the empirical case for the extension and develops its policy implications.

## METHODS

### Study design

We used a concurrent mixed-methods design (Creswell and Plano Clark 2018) in which a household survey (quantitative strand) and key informant interviews and focus group discussions (qualitative strand) were conducted in parallel and given equal priority. The quantitative strand established the prevalence and rank order of enrollment barriers and the socio-demographic predictors of enrollment; the qualitative strand examined the meaning, subtypes and lived experience of institutional distrust. The strands were integrated at the interpretation stage through a joint-display approach (Guetterman et al. 2015). Reporting of the quantitative strand follows the Strengthening the Reporting of Observational Studies in Epidemiology (STROBE) statement for cross-sectional studies (von Elm et al. 2007), and reporting of the mixed-methods design and integration follows the Good Reporting of A Mixed Methods Study (GRAMMS) guideline (O’Cathain et al. 2008); both completed checklists are provided as Supplementary File 2.

### Relationship to a companion analysis

This manuscript reports the barrier analysis from the same cross-sectional study whose health outcome findings are reported in a companion manuscript (Omisore et al., companion manuscript under review). The companion manuscript compares physical, psychological and social health outcomes between insured and uninsured respondents. The present paper addresses a distinct question, namely the barriers to enrollment and the place of institutional trust within the Andersen model, and is the only one of the two to report the forced-choice barrier item, the enrollment regression and the qualitative strand. Overlapping descriptions of the sampling frame and survey procedures are kept consistent with the companion manuscript.

### Study setting

Four Liberian counties were purposively selected to span the country’s geographic, ethnic and infrastructural diversity: Montserrado (urban; includes the capital, Monrovia), Margibi (peri-urban and industrial; rubber and port economy), Nimba (north-eastern; conflict-affected; ethnically heterogeneous) and Grand Gedeh (remote south-eastern; logistically constrained; Ebola-affected). Together these counties account for approximately 45% of Liberia’s population and represent the principal contexts in which any national insurance scheme must operate.

### Quantitative strand: sampling and survey

A stratified sample of 542 adults (aged 18 years or older and principal household decision-makers on health expenditure) was recruited across the four counties, with equal allocation of 271 insured and 271 uninsured respondents to permit direct comparison between groups. Participants were selected through a four-stage design. Within each county, settlements were stratified as urban or rural with proportional allocation; 48 communities (12 per county, balanced between strata) were selected with probability proportional to size from National Population and Housing Census enumeration lists; and households were selected by systematic random sampling from community household registers, with one eligible, consenting adult enrolled per household. Household selection continued within each community until the insured and uninsured quotas were filled; where a stratum could not be filled from households, participants were recruited at health facilities and community gathering points serving the same community and through community health worker referrals, using the same eligibility criteria, consent procedure and instruments. The household stage is therefore best described as stratified quota recruitment within probability-selected communities, and the estimates are not presented as nationally representative. Participant recruitment and data collection took place in May 2026. Sample size was calculated for the insured versus uninsured comparison using the formula for two independent proportions: 246 per group provided 80% power at a two-sided significance level of 0.05 to detect a difference of 12.5 percentage points (60% versus 47.5%), and this was inflated by 10% for non-response to 271 per group (N = 542). The barrier analysis is restricted to the 271 uninsured respondents, a subset of the sample sized for the primary insured versus uninsured comparison; it was not separately powered to detect differences between individual barrier categories or counties. Questionnaires were completed on paper and, for some respondents, through a distributed online link.

The structured questionnaire collected demographic and socioeconomic data, current enrollment status and, for non-enrolled respondents, a forced-choice item on the primary barrier to enrollment ("What is the single most important reason you do not have health insurance?") with six pre-coded response categories adapted from barrier items used in comparable West African community-based health insurance research. Before fieldwork, the questionnaire was reviewed for cultural and linguistic appropriateness by two Liberian public health practitioners, cognitively tested with five community health workers from areas adjacent to the study counties, and piloted with 30 adults in non-study communities in Margibi; cognitive interviews confirmed that the six barrier options were understood and covered the reasons participants would give. Because respondents selected one primary barrier, category percentages sum to 100% and identify the binding constraint each respondent perceived rather than the full set of constraints experienced. The survey did not include multi-item trust scales; the four analytic subtypes of institutional distrust reported below were therefore developed from the qualitative strand, and no quantitative subtype estimates are reported.

### Qualitative strand: sampling and data collection

Sixteen key informant interviews were conducted with purposively selected informants across four stakeholder categories: health insurance administration; county or district health services and health care provision; community health; and national policy and development partners. Informants were selected for their direct experience of insurance design, implementation or uptake. Quotations are attributed using role and county labels only where these do not risk identifying an informant, and the distribution of labels does not imply equal representation of every role in every county.

Four focus group discussions were conducted, one in each of Montserrado, Margibi, Nimba and Grand Gedeh, each designed to include at least four participants drawn from enrolled and non-enrolled community members. The topic guide addressed knowledge of insurance, reasons for enrollment or non-enrollment, experiences with health institutions, expectations of fund stewardship and benefit delivery, and proposed remedies. Key informant interviews were conducted in English using an eight-question semi-structured guide with probes and lasted 45 to 75 minutes; each focus group was led by a trained facilitator with note-taking support. Interviews and discussions were audio-recorded with consent, transcribed, checked against the recordings and field notes, and de-identified before analysis. Reporting of the qualitative strand follows the Consolidated Criteria for Reporting Qualitative Research (COREQ) (Tong et al. 2007); the completed checklist is provided as Supplementary File 1.

### Data analysis

#### Quantitative analysis

Questionnaire data were entered following a structured protocol, with 10% of questionnaires double entered to verify accuracy, and were checked for range and internal consistency. Item-level missingness was below 2% for all items and was not associated with insurance status, county or demographic characteristics, so complete-case analysis was used without imputation. Quantitative analysis was conducted in IBM SPSS Statistics version 26 (IBM Corp., Armonk, NY, USA), with Wilson score 95% confidence intervals for proportions computed in Stata version 17 (StataCorp LLC, College Station, TX, USA). Unadjusted differences between insured and uninsured respondents were examined with Welch’s t-test for age and Pearson’s chi-squared tests for categorical characteristics. Primary barriers to enrollment were described as counts and percentages with Wilson 95% confidence intervals among uninsured respondents, overall and by county, and the distribution of barriers across counties was compared using Pearson’s chi-squared test of independence, with Cramér’s V as a measure of association. Multivariable logistic regression modeled current enrollment (enrolled versus not enrolled) among all 542 respondents, with age (per year), sex, education (no formal education, primary, secondary, tertiary), employment status (unemployed, informally employed, student, formally employed) and county of residence entered simultaneously. Results are reported as adjusted odds ratios (aORs) with 95% confidence intervals (CIs). Because the barrier item was administered only to uninsured respondents and the survey did not include trust scales, institutional distrust was not entered as a regressor; its role was assessed through the barrier distribution and the qualitative strand, and the regression was used to characterize the structural socioeconomic gradient against which perceived barriers operate.

#### Qualitative analysis

Qualitative data were analyzed using reflexive thematic analysis (Braun and Clarke 2006; Braun and Clarke 2019) with a hybrid orientation: Andersen’s predisposing, enabling and need domains provided initial sensitizing concepts, while the subtypes of institutional distrust were developed inductively from the data. Analysis was led by the principal researcher as an interpretive and reflexive process that moved through familiarization, systematic coding, generation and review of candidate themes, and definition and naming of final themes. Consistent with this approach, statistical intercoder agreement was not used as a criterion of analytic quality. Rigor was supported through sustained engagement with the transcripts, reflexive memo-writing, transparent documentation of analytic decisions, deliberate attention to divergent and disconfirming accounts, and the use of illustrative participant quotations.

#### Integration

The strands were integrated at the interpretation stage. Survey barrier rankings were set against the qualitative themes in a joint display (presented in Results) that links each distrust subtype to its defining concern, representative quotations and the corresponding policy lever, and the resulting meta-inferences informed the proposed extension of the Andersen model, which is set out at the end of Results.

### Ethical considerations

Ethical approval was granted by the Research Ethics Committee of the University of Port Harcourt (December 2024; reference UPH/CEREMAD/REC/MM105/030). A letter of local institutional support for data collection in Liberia was obtained (May 2026). Written informed consent was obtained from all participants before data collection, including specific consent for audio recording. Participation was voluntary, participants received no compensation beyond reimbursement of transport costs, and they could withdraw at any time without consequence. Transcripts and survey records were de-identified before analysis.

### Patient and public involvement

Community leaders in each study county were consulted during instrument design and advised on language, the framing of trust-related response options and culturally appropriate phrasing of sensitive questions. Preliminary findings were presented back to community focus groups in each county before manuscript preparation, and their reactions informed the policy implications set out in the Discussion.

## RESULTS

### Sample characteristics

Of approximately 600 eligible adults approached, 58 declined (response rate 90.3%); all 542 enrolled participants were included in the analysis, of whom 271 (50.0%) were currently enrolled in the national insurance scheme and 271 (50.0%) were uninsured. The insured and uninsured strata were of equal size by design, permitting direct comparison within a single multivariable model. Insured respondents were older than uninsured respondents (mean 44.6 years, SD 11.1, against 39.0 years, SD 12.5; p < 0.001). The groups differed markedly in education: 31.7% of uninsured respondents had no formal education and 42.4% primary education only, compared with 11.1% and 20.3% of insured respondents (χ² = 100.79; p < 0.001; Cramér’s V = 0.431). They also differed in employment: 43.5% of uninsured respondents were unemployed and 10.3% formally employed, compared with 23.2% and 45.8% of insured respondents (χ² = 86.56; p < 0.001; V = 0.400). The groups did not differ by sex (χ² = 0.36; p = 0.55) or county of residence (χ² = 3.14; p = 0.37). These unadjusted differences foreshadow the socioeconomic gradient identified in the multivariable analysis.

### Primary barriers to enrollment among the uninsured

Among uninsured respondents (n = 271), institutional distrust was the most frequently reported primary barrier, named by 57 (21.0%; 95% CI 16.6 to 26.3), narrowly ahead of premium cost (49, 18.1%; 95% CI 14.0 to 23.1) and inaccessibility of services (47, 17.3%; 95% CI 13.3 to 22.3) (Table 1). These three leading barriers together accounted for 56.5% of primary reasons for non-enrollment. The remaining responses were that insurance was not necessary (45, 16.6%), not knowing how to enroll (38, 14.0%) and a preference for paying out of pocket (35, 12.9%).

**Table 1.** Distribution of self-reported primary barriers to enrollment among uninsured respondents, overall and by county (n = 271)

| **Barrier category** | **Overall, n (%)** | **Montserrado, %** | **Margibi, %** | **Nimba, %** | **Grand Gedeh, %** |
|---|---|---|---|---|---|
| Do not trust the insurance scheme | 57 (21.0) | 16.9 | 18.3 | 30.8 | 18.8 |
| Cost of premium is too high | 49 (18.1) | 23.9 | 21.1 | 15.4 | 10.9 |
| Services are too far away | 47 (17.3) | 16.9 | 22.5 | 15.4 | 14.1 |
| Do not think it is necessary | 45 (16.6) | 15.5 | 12.7 | 13.8 | 25.0 |
| Do not know how to enroll | 38 (14.0) | 15.5 | 12.7 | 15.4 | 12.5 |
| Prefer to pay out of pocket | 35 (12.9) | 11.3 | 12.7 | 9.2 | 18.8 |
| Total | 271 (100.0) | 100.0 (n = 71) | 100.0 (n = 71) | 100.0 (n = 65) | 100.0 (n = 64) |

Note: Forced-choice item; each respondent selected one primary barrier. Counts are among the 271 uninsured respondents. County percentages are within-county (denominators: Montserrado 71, Margibi 71, Nimba 65, Grand Gedeh 64). The barrier distribution did not differ significantly by county (Pearson χ<sup>2</sup> = 15.99, df = 15; p = 0.38; Cramér's V = 0.140; minimum expected cell count 8.27). Percentages may not sum to exactly 100 because of rounding.

The barrier distribution did not differ significantly across the four counties (χ² = 15.99, df = 15; p = 0.38; Cramér’s V = 0.140), indicating that the ordering of constraints was broadly consistent across the study counties rather than confined to a single county. Two descriptive patterns are nonetheless notable. Distrust was most frequently cited in Nimba (30.8%), the county with the most intense exposure to civil conflict, whereas in Grand Gedeh the most frequent response was that insurance is unnecessary (25.0%). Because each respondent selected a single primary barrier, these figures identify the binding constraint each respondent perceived rather than every constraint faced; the qualitative strand indicates that distrust frequently coexists with cost and access constraints rather than substituting for them.

### Subtypes of institutional distrust: qualitative findings

Qualitative analysis organized accounts of institutional distrust into four analytic subtypes, each with a distinct defining concern and a corresponding policy lever (Table 2). Three subtypes were directly illustrated by retained quotations; evidence for provider-quality distrust overlapped with broader distrust of the state health sector and this subtype should therefore be regarded as provisional. Together, the subtypes show that the single survey category "do not trust the insurance scheme" conceals several different expectations of failure grounded in institutional experience.

**Table 2.** Institutional distrust subtypes: concerns, quotations and policy levers.

| **Subtype** | **Defining concern** | **Representative quotation** | **Policy lever** |
|---|---|---|---|
| Fund-stewardship distrust | Premiums will be misappropriated or diverted before reaching the benefit pool | "People lost everything. So when you come and say put your money in insurance, my mind goes to that experience." (Margibi FGD participant who lost savings when a financial institution collapsed) | Independent audit; public reporting; ring-fenced accounts |
| Benefit-delivery distrust | Promised services will not be available when claimed (stockouts, denials) | "We have cases where claims were submitted months ago and are still pending. When communities hear about this, it doesn't matter how well we explain our scheme, the word gets around that insurance doesn't pay." (Montserrado insurance manager, KII) | Supply-chain reliability; published benefit guarantees; grievance redress |
| Provider-quality distrust | Insurance-affiliated facilities will not deliver care of acceptable quality to enrolled patients | Provisional analytic subtype: no quotation specific to individual provider quality was retained; the illustrative Grand Gedeh account is reported under generalized-state distrust. | Provider payment design; quality monitoring; complaint channels |
| Generalized-state distrust | Diffuse expectation that state institutions act in bad faith | "We have seen what governments do here in Nimba. They make promises during campaigns and then forget us after. We have been burned too many times. This insurance is another promise from people who may not be here to keep it." (Nimba FGD participant)<br>"During Ebola, people saw that the government system failed to protect health workers, who themselves became victims. When we now come asking people to invest their money in a health insurance scheme managed by that same sector, we inherit the credibility deficit that the EVD response created." (Grand Gedeh district health officer, KII) | Long-horizon trust-building; multi-stakeholder governance; track-record accumulation |

Note: Quotations were selected for thematic representativeness across the qualitative corpus. The Grand Gedeh quotation links expectations of care under insurance to the health sector's wider credibility after Ebola and is interpreted as evidence about institutional trust in the sector rather than the clinical quality of individual providers. EVD = Ebola virus disease; FGD = focus group discussion; KII = key informant interview.

#### Fund-stewardship distrust

Fund-stewardship distrust was the expectation that premiums would be misappropriated or diverted before reaching the benefit pool. It could be anchored in personal experience of earlier institutional failures involving household savings. As one Margibi focus group participant, who had lost savings when a financial institution collapsed, explained: "People lost everything. So when you come and say put your money in insurance, my mind goes to that experience." Here, reluctance to enroll reflected a reasoned transfer of experience from one failed custodian of household money to another prospective one.

#### Benefit-delivery distrust

Benefit-delivery distrust concerned whether promised services would be available when claimed. Informants described how delays in the claims process, once known in communities, undermined outreach regardless of its quality: "We have cases where claims were submitted months ago and are still pending. When communities hear about this, it doesn’t matter how well we explain our scheme, the word gets around that insurance doesn’t pay." (Montserrado insurance manager, key informant interview). This account illustrates how administrative performance is converted into community reputation, and why information campaigns alone cannot offset visible delivery failures.

#### Provider-quality distrust

Provider-quality distrust concerned whether insurance-affiliated facilities would deliver acceptable care to enrolled patients. Participants’ accounts of this concern were closely bound up with the health sector’s wider track record rather than with the clinical skills of individual providers, so the most illustrative quotation, from Grand Gedeh, is reported under generalized-state distrust below.

#### Generalized-state distrust

Generalized-state distrust was the diffuse expectation that state institutions act in bad faith. A participant from Nimba, the county where distrust was most often named as the primary survey barrier, expressed it directly: "We have seen what governments do here in Nimba. They make promises during campaigns and then forget us after. We have been burned too many times. This insurance is another promise from people who may not be here to keep it." (Nimba focus group participant). In such accounts, the insurance scheme was not evaluated on its own design but as one more instance of a long series of state commitments, which is why this subtype is likely to be the most resistant to short-term remedy. The same deficit extended to the health sector itself. A district health officer in Grand Gedeh traced it to the Ebola response: "During Ebola, people saw that the government system failed to protect health workers, who themselves became victims. When we now come asking people to invest their money in a health insurance scheme managed by that same sector, we inherit the credibility deficit that the EVD response created." (Grand Gedeh district health officer, key informant interview). Accounts of this kind show that judgments about the care an insured patient will receive are shaped by the state health sector’s wider record, and we interpret them as evidence about institutional trust rather than about the clinical quality of individual providers.

### Socio-demographic predictors of enrollment

Multivariable logistic regression with current enrollment as the outcome identified socioeconomic status as the strongest structural predictor (Table 3). Compared with unemployed respondents, those in formal employment had nearly nine times the odds of enrollment (aOR 8.78, 95% CI 4.90 to 15.76; p < 0.001). Compared with respondents with no formal education, the odds of enrollment were higher among those with tertiary education (aOR 8.17, 95% CI 4.12 to 16.20; p < 0.001) and secondary education (aOR 7.36, 95% CI 3.98 to 13.63; p < 0.001), whereas primary education was not associated with enrollment (aOR 1.40, 95% CI 0.77 to 2.55; p = 0.27). Each additional year of age was associated with a small increase in the odds of enrollment (aOR 1.05, 95% CI 1.03 to 1.07; p < 0.001). Informal employment (aOR 1.35, 95% CI 0.81 to 2.24; p = 0.25) and student status (aOR 1.14, 95% CI 0.53 to 2.45; p = 0.74) were not significantly associated with enrollment, nor were female sex (aOR 1.20, 95% CI 0.79 to 1.82; p = 0.40) or county of residence (all p ≥ 0.29).

**Table 3.** Multivariable logistic regression of socio-demographic predictors of health insurance enrollment (N = 542)

| **Predictor** | **Adjusted OR** | **95% CI** | **p-value** |
|---|---|---|---|
| Age (per year) | 1.05 | 1.03 to 1.07 | <0.001 |
| Sex (female; male = reference) | 1.20 | 0.79 to 1.82 | 0.40 |
| *Education (no formal education = reference)* |  |  |  |
| Primary | 1.40 | 0.77 to 2.55 | 0.27 |
| Secondary | 7.36 | 3.98 to 13.63 | <0.001 |
| Tertiary | 8.17 | 4.12 to 16.20 | <0.001 |
| *Employment (unemployed = reference)* |  |  |  |
| Informally employed | 1.35 | 0.81 to 2.24 | 0.25 |
| Student | 1.14 | 0.53 to 2.45 | 0.74 |
| Formally employed | 8.78 | 4.90 to 15.76 | <0.001 |
| *County (Grand Gedeh = reference)* |  |  |  |
| Montserrado | 0.72 | 0.39 to 1.33 | 0.29 |
| Margibi | 1.25 | 0.70 to 2.23 | 0.45 |
| Nimba | 1.12 | 0.62 to 2.00 | 0.71 |

Note: Outcome is current enrollment (1) versus non-enrollment (0); an adjusted odds ratio (aOR) greater than 1 indicates higher odds of enrollment. All predictors were entered simultaneously. Reference categories: male; no formal education; unemployed; Grand Gedeh. CI = confidence interval.

Read together with the barrier data, these results show two complementary layers. The regression identifies who is structurally positioned to enroll, chiefly the formally employed and those with at least secondary education, while the survey and qualitative strands converge on institutional distrust as the barrier the uninsured themselves most often name. The absence of an association with informal employment is particularly relevant for policy, because informal-sector households are the population that voluntary contributory schemes most need to reach and the population for whom trust in the scheme matters most when deciding whether to pay.

### Locating institutional trust in the Andersen framework

Table 4 sets out the proposed extension, juxtaposing the conventional Andersen enabling-resources structure with the four-subcategory revision. The change is additive: predisposing and need factors are unaffected; the financial, organizational and social-support subcategories of enabling resources are retained unchanged; and institutional-trust resources are added as a fourth subcategory comprising the four subtypes identified above. Measurement implications follow directly: each trust subtype becomes a candidate variable for Andersen-framed studies, to be measured alongside existing income, proximity and social-support measures.

**Table 4.** Proposed Andersen Behavioral Model extension: enabling-resource subcategories.

| **Subcategory** | **Definition** | **Standard measurement examples** | **Status in proposal** |
|---|---|---|---|
| Financial resources | Personal and household means to pay for or finance care | Income; insurance status; capacity for out-of-pocket expenditure | Retained (Andersen) |
| Organizational resources | System-level capacity facilitating access | Provider availability; geographic proximity; regular source of care | Retained (Andersen) |
| Social-support resources | Family, kin and community networks supporting service use | Household composition; community network density; caregiver availability | Retained (Andersen) |
| Institutional-trust resources | Credible expectation that fund custodians, providers and the state will deliver promised services | Fund-stewardship trust; benefit-delivery trust; provider-quality trust; generalized-state trust | Proposed addition |

Note: The proposed addition preserves the model's existing structure and feedback logic; it adds a fourth parallel subcategory to enabling resources without modifying predisposing characteristics or need factors. Measurement examples for institutional-trust resources derive from the qualitative findings reported in Table 2 and are proposed for operationalization in future survey instruments; they were not measured as scales in the present survey.

## DISCUSSION

### Principal findings

Three findings stand out. First, institutional distrust was the most frequently reported primary barrier to health insurance enrollment among uninsured adults in post-conflict Liberia, narrowly ahead of premium cost and inaccessibility, and the distribution of barriers did not differ significantly across four counties that differ markedly in urbanization, conflict exposure and remoteness. Second, distrust is not monolithic: the analysis generated four distinguishable subtypes with different policy implications, although the provider-quality subtype requires further direct validation. Third, distrust operates alongside, rather than in place of, a steep socioeconomic gradient: formal employment and secondary or tertiary education were the strongest independent predictors of enrollment. The two sets of findings are complementary. Socioeconomic status describes who is structurally able to enroll; institutional distrust describes why many of those who remain outside the scheme say they have not joined.

### Trust as an enabling resource: theoretical implications

These findings support the proposed extension of the Andersen Behavioral Model. The conventional treatment of trust as a predisposing belief understates its role in fragile-state contexts in three ways that the data make visible. It conflates a contingent assessment of institutional behavior with a stable individual attribute, whereas participants consistently linked their distrust to specific institutional events: a collapsed financial institution, delayed claims, the health sector’s performance during Ebola and unkept political promises. It implies that distrust can be remedied through information, whereas informants described communities discounting well-explained schemes once delivery failures became known. And it separates trust analytically from the structural resources it most closely resembles, whereas in participants’ accounts a credible expectation of institutional performance functioned as a precondition for paying into a scheme, in much the way that money or transport functions as a precondition for attending a facility.

The proposed reframing is theoretically conservative. It does not modify the model’s underlying causal logic, displaces no existing category and remains testable with the model’s standard empirical apparatus. It is nonetheless consequential: it implies that Andersen-framed studies of service use in fragile states that omit institutional-trust variables risk misspecification, and that interventions targeting only the conventional enabling categories may systematically underperform. The regression results sharpen rather than weaken this argument. Because the strongest structural predictors (education and formal employment) cannot be changed quickly by an insurance agency, institutional trust is among the few enabling resources that scheme managers can influence directly through their own conduct.

### Comparison with prior literature

The prominence of distrust among enrollment barriers in a post-conflict African setting is consistent with evidence from Sierra Leone (Pieterse and Lodge 2015) and the Democratic Republic of Congo (Vinck et al. 2019). This study adds, first, a formal disaggregation of distrust into subtypes with distinct policy implications and, second, the theoretical integration of trust into a widely used service-use framework rather than its treatment as a standalone construct.

Prior work on trust in health systems has tended either to treat trust as a unitary attitudinal variable (Gilson 2003; Ozawa and Sripad 2013) or to develop trust-specific frameworks that do not connect to mainstream service-use models (Gille et al. 2021). The extension proposed here is designed to bridge these traditions. The coexistence of distrust with a strong education and employment gradient also echoes the broader UHC literature, which emphasizes that voluntary contributory schemes struggle to reach informal-sector populations (Carrin et al. 2008; Kutzin 2013); our findings suggest that institutional distrust is one reason that struggle persists even where premiums are modest.

### Policy implications

If institutional trust is an enabling resource, trust-building should appear as a programmatic and budgeted line item in enrollment policy rather than as a soft adjunct. Four implications follow, mapping to the four subtypes identified.

Fund-stewardship distrust is likely to respond primarily to demonstrable financial accountability: ring-fenced accounts, independent audit, public reporting of premium collection and benefit payout, and visible separation of the insurance pool from general government revenue. Because these measures are within the direct control of the scheme, they are plausible early-moving levers whose effect on stewardship trust could be tracked over the first one to two years of implementation.

Benefit-delivery distrust is likely to respond to reliable supply and credible grievance redress: stockouts should be tracked and published, claim-processing times and denial rates made transparent, and accessible mechanisms established for resolving delayed or denied claims. Where benefit guarantees are not stated publicly and tracked, skepticism among potential enrollees is a reasonable response to the information available to them.

Provider-quality distrust is likely to respond to provider-payment design and quality monitoring. Payment structures that create incentives to under-serve enrolled patients should be redesigned, quality indicators tracked separately for enrolled and out-of-pocket patients, and complaint channels made accessible and demonstrably responsive. Because participants linked expectations of care to the health sector’s wider record, visible improvements in the protection and support of health workers may also contribute.

Generalized-state distrust is likely to be the most resistant to short-term remedy. It accumulates only through demonstrated institutional reliability across multiple programs over time. Multi-stakeholder governance, with donor, civil society and faith-based representation on oversight bodies, can partly substitute for state credibility during the rebuilding phase, but the principal lever is a sustained track record. In parallel, measures that lower financial barriers for informal-sector and less-educated households, such as targeted premium subsidies and flexible contribution schedules, remain essential, because trust-building alone cannot offset the socioeconomic gradient identified in the regression.

### Implications for research

Three research directions follow. First, a multi-item instrument measuring the four trust subtypes should be developed and psychometrically validated in Liberia and other fragile-state settings, with attention to cross-cultural translation and equivalence; this would allow institutional trust to be entered directly into enrollment models alongside socioeconomic variables. Second, longitudinal application of the extended Andersen model should test whether changes in trust subtypes predict changes in enrollment over time. Third, intervention studies should pair trust-building components (audit, grievance redress, provider-payment reform) with conventional enrollment interventions to estimate the effect on enrollment attributable to trust.

### Strengths and limitations

The study has several strengths. The concurrent mixed-methods design allowed direct triangulation between the prevalence of perceived barriers and their meaning. The four-county sample spans substantial contextual variation within Liberia, from the capital to remote, conflict-affected and Ebola-affected counties, and the absence of significant county differences in the barrier distribution suggests that the pattern was not confined to one study county. The equal allocation of insured and uninsured respondents supported direct comparison of insured and uninsured adults within a single multivariable model of enrollment. The response rate was high (90.3%), item-level missingness was below 2%, and the barrier item was cognitively tested and piloted before fieldwork. Finally, the theoretical extension is grounded in primary data rather than literature synthesis alone, and preliminary findings were presented back to participating communities.

Several limitations should be noted. First, the findings come from Liberia and may not generalize to non-fragile sub-Saharan settings where trust dynamics differ. Second, institutional trust was captured in the survey through a single forced-choice barrier category rather than a validated multi-item scale, so trust could not be entered into the enrollment regression and the four analytic subtypes require independent validation, particularly the provisional provider-quality subtype. Third, because the barrier item recorded only one primary reason, the prevalence of distrust as a contributing barrier is likely to be higher than its prevalence as the primary barrier, while the small margin between distrust (21.0%) and cost (18.1%) means that the two should be regarded as comparably important. Fourth, the cross-sectional design constrains causal inference, and longitudinal data would strengthen the case. Fifth, the proposed model extension is at this stage a hypothesis with empirical motivation; full validation requires application in additional fragile-state settings.

The mixed-mode survey administration also has limitations. Participation through a distributed online link may have introduced differential access associated with digital literacy and connectivity, as well as self-selection, and responses collected online and on paper may be subject to mode effects. Supplementary recruitment at facilities and community sites, used where households could not fill an insurance stratum, may over-represent adults in recent contact with health services, and enrolling any eligible adult in each household rather than applying a within-household selection grid may over-represent adults who were at home or had access to a device. Because communities were the primary sampling units and the analyses did not adjust for clustering by community, the confidence intervals may be somewhat narrower than design-based estimates would be. Some qualitative reporting details recommended by COREQ, such as focus group size and focus group duration, were not systematically recorded and are reported as not available. These considerations qualify the representativeness of the sample where access to, or uptake of, the online option differed among eligible participants.

## CONCLUSIONS

In post-conflict Liberia, institutional distrust emerged as the most frequently reported barrier to health insurance enrollment among the uninsured, narrowly ahead of cost (21.0% against 18.1%), and it operated alongside socioeconomic status and access as a distinct and policy-addressable constraint that conventional affordability measures do not reach. Socioeconomic status, particularly formal employment and secondary or tertiary education, remained the strongest structural predictor of enrollment. The analysis generated four distrust subtypes with distinct policy implications; three were directly anchored in retained participant quotations, while the provider-quality subtype remains provisional and requires further validation. The Andersen Behavioral Model, the most widely applied framework in health services research, conventionally treats trust as a predisposing belief; the evidence presented here suggests that in fragile-state settings it functions instead as an enabling resource on a par with financial, organizational and social-support resources. We have therefore proposed a conservative extension of the model that adds institutional trust as a fourth subcategory of enabling resources, with four analytic subtypes for further testing. The policy implication is direct: in fragile-state contexts, trust-building is health-system infrastructure that must accompany, not replace, measures that improve financial access, and enrollment policy that omits it is likely to underperform. The extension remains a hypothesis pending wider validation, but the survey, key informant and focus group evidence motivating it converge, and the cost of misspecification, persistent under-enrollment despite well-designed schemes, is too well documented to leave the framework unrevised.

## DECLARATIONS

### Ethics approval

Ethical approval was granted by the Research Ethics Committee of the University of Port Harcourt (December 2024; reference UPH/CEREMAD/REC/MM105/030). A letter of local institutional support for data collection in Liberia was obtained (May 2026). Written informed consent was obtained from all participants.

### Consent to participate

Written informed consent was obtained from all survey respondents, key informants and focus group participants before data collection, including specific consent for audio recording. Participation was voluntary and participants could withdraw at any time without consequence.

### Consent for publication and preprint posting: All authors have read and approved the manuscript and consent to its posting as a preprint

The manuscript contains no individually identifiable data; quotations are attributed only by role, data collection method and county.

### Data availability

De-identified data supporting the findings are available from the corresponding author on reasonable request, subject to participant confidentiality.

### Author contributions (CRediT)

O.K.O.: Conceptualization, Methodology, Investigation, Formal analysis, Writing (original draft). B.W.: Resources, Investigation (facilitation of field access and data collection in Liberia), Writing (review and editing). V.A.: Supervision, Validation, Writing (review and editing). D.O.: Supervision, Methodology, Writing (review and editing). All authors approved the final version.

## Acknowledgments

We thank the community leaders, key informants and focus group participants in Montserrado, Margibi, Nimba and Grand Gedeh counties for their time and candor. We acknowledge logistical support from Hill-City University, Liberia.

## Supplementary material

Supplementary File 1: completed Consolidated Criteria for Reporting Qualitative Research (COREQ) checklist, appended to this manuscript, indicating where each item is reported or marking it as partially reported or not reported. Supplementary File 2: completed STROBE checklist for cross-sectional studies and GRAMMS checklist for mixed-methods studies, appended to this manuscript, indicating where each item is reported or marking it as partially reported or not reported.

---

**Supplementary File 1. Consolidated Criteria for Reporting Qualitative Research (COREQ) checklist**

Institutional Distrust as the Most Frequently Reported Barrier to Health Insurance Enrollment in Post-Conflict Liberia: A Mixed-Methods Study Extending the Andersen Behavioral Model with Trust as a Fourth Enabling Resource

COREQ 32-item checklist for interviews and focus groups (Tong et al. 2007)

| No. | Item | Where reported in this manuscript |
|---|---|---|
| **Domain 1: Research team and reflexivity** |  |  |
| ***Personal characteristics*** |  |  |
| 1 | Interviewer or facilitator: Which author(s) conducted the interview or focus group? | Methods, Qualitative strand: sampling and data collection (interviews and focus groups conducted by the principal researcher, the first author, with assigned trained field and data officers; focus groups led by a trained facilitator with note-taking support) |
| 2 | Credentials: What were the researcher's credentials? (e.g., PhD, MD) | Partially reported: Title page (qualifications of the principal researcher, the first author, who led data collection); credentials of the assigned field and data officers are not stated |
| 3 | Occupation: What was their occupation at the time of the study? | Title page; Reflexivity statement (first author a doctoral researcher at the University of Port Harcourt; co-authors are public health and health systems researchers at Nigerian institutions and in Liberia) |
| 4 | Gender: Was the researcher male or female? | Not reported |
| 5 | Experience and training: What experience or training did the researcher have? | Partially reported: Methods, Qualitative strand: sampling and data collection (data collected by the principal researcher, a doctoral researcher, with assigned trained field and data officers; focus groups led by a trained facilitator); the content and duration of training are not described |
| ***Relationship with participants*** |  |  |
| 6 | Relationship established: Was a relationship established prior to study commencement? | Not reported |
| 7 | Participant knowledge of the interviewer: What did the participants know about the researcher? (e.g., personal goals, reasons for doing the research) | Not reported |
| 8 | Interviewer characteristics: What characteristics were reported about the interviewer or facilitator? (e.g., bias, assumptions, reasons and interests in the research topic) | Title page, reflexivity statement |
| **Domain 2: Study design** |  |  |
| ***Theoretical framework*** |  |  |
| 9 | Methodological orientation and theory: What methodological orientation was stated to underpin the study? (e.g., grounded theory, discourse analysis, ethnography, phenomenology, content analysis) | Methods, Qualitative analysis (reflexive thematic analysis, hybrid orientation) |
| ***Participant selection*** |  |  |
| 10 | Sampling: How were participants selected? (e.g., purposive, convenience, consecutive, snowball) | Methods, Qualitative strand: sampling and data collection (purposive selection of key informants for direct experience of insurance design, implementation or uptake across four stakeholder categories; focus group participants drawn from enrolled and non-enrolled community members) |
| 11 | Method of approach: How were participants approached? (e.g., face-to-face, telephone, mail, email) | Partially reported: Methods, Quantitative strand: sampling and survey; Discussion, Strengths and limitations (mixed approach for survey participants: household visits, facility, community-site and CHW referral, and a distributed online link); how key informants and focus group participants were approached is not stated |
| 12 | Sample size: How many participants were in the study? | Partially reported: Methods, Qualitative strand: sampling and data collection (16 key informant interviews; four focus group discussions, one per county, each designed to include at least four participants, giving a planned minimum of 32 qualitative participants); achieved focus group sizes are not stated |
| 13 | Non-participation: How many people refused to participate or dropped out? Reasons? | Not reported |
| ***Setting*** |  |  |
| 14 | Setting of data collection: Where were the data collected? (e.g., home, clinic, workplace) | Partially reported: Methods, Study setting and Qualitative strand: sampling and data collection (one focus group in each of Montserrado, Margibi, Nimba and Grand Gedeh); specific venues of interviews and focus groups are not stated |
| 15 | Presence of non-participants: Was anyone else present besides the participants and researchers? | Not reported |
| 16 | Description of sample: What are the important characteristics of the sample? (e.g., demographic data, date) | Methods, Qualitative strand: sampling and data collection (key informants from health insurance administration, county or district health services and care provision, community health, and national policy; focus groups of enrolled and non-enrolled community members in four counties); Methods, Quantitative strand: sampling and survey (data collection period, May 2026) |
| ***Data collection*** |  |  |
| 17 | Interview guide: Were questions, prompts and guides provided by the authors? Was it pilot tested? | Partially reported: Methods, Qualitative strand: sampling and data collection (eight-question semi-structured guide with probes; topic guide content); pilot testing of the qualitative guide is not described |
| 18 | Repeat interviews: Were repeat interviews carried out? If yes, how many? | Not reported |
| 19 | Audio or visual recording: Did the research use audio or visual recording to collect the data? | Methods, Qualitative strand: sampling and data collection (interviews and focus groups audio-recorded with consent; transcripts checked against recordings and field notes, and de-identified) |
| 20 | Field notes: Were field notes made during and/or after the interview or focus group? | Methods, Qualitative strand: sampling and data collection (field notes taken by note-taking support during focus groups) |
| 21 | Duration: What was the duration of the interviews or focus group? | Partially reported: Methods, Qualitative strand: sampling and data collection (key informant interviews 45 to 75 minutes); focus group duration is not stated |
| 22 | Data saturation: Was data saturation discussed? | Not reported |
| 23 | Transcripts returned: Were transcripts returned to participants for comment and/or correction? | Not reported |
| **Domain 3: Analysis and findings** |  |  |
| ***Data analysis*** |  |  |
| 24 | Number of data coders: How many data coders coded the data? | Methods, Qualitative analysis (one coder: analysis led by the principal researcher; consistent with reflexive thematic analysis, intercoder agreement was not used as a quality criterion) |
| 25 | Description of the coding tree: Did authors provide a description of the coding tree? | Partially reported: Methods, Qualitative analysis (Andersen's predisposing, enabling and need domains as initial sensitizing concepts; phases from familiarization and systematic coding to definition and naming of themes); Results, Subtypes of institutional distrust (four inductive subtypes); the full coding tree is not reported |
| 26 | Derivation of themes: Were themes identified in advance or derived from the data? | Methods, Qualitative analysis (hybrid: Andersen domains provided initial sensitizing concepts; subtypes of institutional distrust were derived inductively from the data) |
| 27 | Software: What software, if applicable, was used to manage the data? | Not reported |
| ***Reporting*** |  |  |
| 28 | Participant checking: Did participants provide feedback on the findings? | Methods, Patient and public involvement (findings presented back to community focus groups) |
| 29 | Quotations presented: Were participant quotations presented to illustrate the themes or findings? Was each quotation identified? (e.g., participant number) | Results, Subtypes of institutional distrust and Table 2; quotations are identified by role, data collection method and county, with labels used only where they do not risk identifying an informant (Methods, Qualitative strand; Declarations, Consent for publication) |
| 30 | Data and findings consistent: Was there consistency between the data presented and the findings? | Results; Table 2; Methods, Qualitative analysis (rigor supported by sustained engagement with transcripts, reflexive memo-writing and documented analytic decisions) |
| 31 | Clarity of major themes: Were major themes clearly presented in the findings? | Results, Subtypes of institutional distrust |
| 32 | Clarity of minor themes: Is there a description of diverse cases or discussion of minor themes? | Results, Subtypes of institutional distrust; Methods, Qualitative analysis (deliberate attention to divergent and disconfirming accounts) |

Item wording follows the COREQ checklist (Tong et al. 2007). Locations refer to section headings and tables in the main manuscript. "Partially reported" indicates that the item is addressed in part, with the missing element stated; "Not reported" indicates that the item is not described in the manuscript.

**Supplementary File 2. STROBE checklist for cross-sectional studies and Good Reporting of A Mixed Methods Study (GRAMMS) checklist**

Institutional Distrust as the Most Frequently Reported Barrier to Health Insurance Enrollment in Post-Conflict Liberia: A Mixed-Methods Study Extending the Andersen Behavioral Model with Trust as a Fourth Enabling Resource

**Table A.** STROBE checklist for cross-sectional studies (von Elm et al. 2007)

| **No.** | **Item** | **Where reported in this manuscript** |
|---|---|---|
| **Title and abstract** |  |  |
| 1a | Indicate the study's design with a commonly used term in the title or the abstract | Partially reported: the title identifies a mixed-methods study and the Abstract describes a concurrent mixed-methods design with a stratified survey; the term "cross-sectional" is used in Methods (Relationship to a companion analysis) but not in the title or Abstract |
| 1b | Provide in the abstract an informative and balanced summary of what was done and what was found | Abstract |
| **Introduction** |  |  |
| 2 | Background/rationale: explain the scientific background and rationale for the investigation being reported | Introduction; Conceptual framework (The Andersen Behavioral Model: structure and limits; Why institutional trust is enabling, not predisposing) |
| 3 | Objectives: state specific objectives, including any prespecified hypotheses | Introduction (final two paragraphs: aim and three contributions); Conceptual framework (Proposed extension: trust as a fourth enabling resource). No prespecified statistical hypotheses are stated |
| **Methods** |  |  |
| 4 | Study design: present key elements of study design early in the paper | Methods, Study design; Relationship to a companion analysis |
| 5 | Setting: describe the setting, locations, and relevant dates, including periods of recruitment, exposure, follow-up, and data collection | Methods, Study setting (four counties) and Quantitative strand: sampling and survey (recruitment and data collection in May 2026; recruitment sources) |
| 6a | Participants: give the eligibility criteria, and the sources and methods of selection of participants | Methods, Quantitative strand: sampling and survey (adults aged 18 years or older who were principal household decision-makers on health expenditure; four-stage design with urban/rural stratification and proportional allocation; 48 communities, 12 per county, selected by probability proportional to size from census enumeration lists; systematic random household selection from community registers; one adult per household; quota fill with facility, community-site and community health worker referral top-up) |
| 7 | Variables: clearly define all outcomes, exposures, predictors, potential confounders, and effect modifiers. Give diagnostic criteria, if applicable | Partially reported: Methods, Quantitative strand: sampling and survey (forced-choice primary barrier item with six categories) and Quantitative analysis (outcome: current enrollment; predictors: age, sex, education, employment status, county). Potential confounders and effect modifiers are not separately specified |
| 8 | Data sources/measurement: for each variable of interest, give sources of data and details of methods of assessment (measurement). Describe comparability of assessment methods if there is more than one group | Partially reported: Methods, Quantitative strand: sampling and survey (structured questionnaire, paper and online administration; barrier item adapted from West African community-based health insurance research, with expert review by two Liberian public health practitioners, cognitive testing with five community health workers and a pilot with 30 adults in non-study Margibi communities); Patient and public involvement (community input on instrument wording). Comparability of paper and online modes was not assessed (noted in Strengths and limitations) |
| 9 | Bias: describe any efforts to address potential sources of bias | Methods, Quantitative strand: sampling and survey (probability proportional to size selection of communities and systematic random household selection) and Quantitative analysis (10% double entry; range and consistency checks); Discussion, Strengths and limitations (facility and community-site recruitment may over-represent recent service contact; any-adult household selection may over-represent at-home or device access; mixed-mode effects; single primary barrier response). Analyses were not adjusted for clustering by community (Discussion, Strengths and limitations) |
| 10 | Study size: explain how the study size was arrived at | Methods, Quantitative strand: sampling and survey (two independent proportions, 60% versus 47.5%, a 12.5 percentage point difference, at 80% power and alpha of 0.05: 246 per group plus 10% non-response, giving 271 per group and N = 542; the barrier analysis of the 271 uninsured respondents was not separately powered to detect differences between barrier categories or counties) |
| 11 | Quantitative variables: explain how quantitative variables were handled in the analyses. If applicable, describe which groupings were chosen and why | Methods, Quantitative analysis (age per year; education and employment categories listed); Table 3. Reasons for the chosen groupings are not stated |
| 12a | Statistical methods: describe all statistical methods, including those used to control for confounding | Methods, Quantitative analysis (Welch's t-test for age and Pearson's chi-squared test for categorical variables in unadjusted comparisons; Wilson score 95% CIs; Pearson's chi-squared test and Cramér's V; multivariable logistic regression with simultaneous entry of covariates; IBM SPSS Statistics version 26, with Wilson 95% CIs in Stata version 17) |
| 12b | Describe any methods used to examine subgroups and interactions | Partially reported: Methods, Quantitative analysis (barrier distribution compared across counties). No interaction terms were examined |
| 12c | Explain how missing data were addressed | Methods, Quantitative analysis (item-level missingness below 2%, not associated with insurance status, county or demographics; complete-case analysis with no imputation) |
| 12d | If applicable, describe analytical methods taking account of sampling strategy | Partially reported: the four-stage stratified design, with equal allocation by insurance status (271 insured, 271 uninsured) and proportional urban/rural allocation within counties, is described (Quantitative strand: sampling and survey); no weighting or adjustment for clustering is described |
| 12e | Describe any sensitivity analyses | Not reported |
| **Results** |  |  |
| 13a | Participants: report numbers of individuals at each stage of study (e.g., numbers potentially eligible, examined for eligibility, confirmed eligible, included in the study, completing follow-up, and analyzed) | Results, Sample characteristics (about 600 eligible adults approached, 58 declined, response rate 90.3%; 542 recruited and analyzed: 271 insured, 271 uninsured); Table 1 (271 uninsured by county); Table 3 (N = 542) |
| 13b | Give reasons for non-participation at each stage | Not reported |
| 13c | Consider use of a flow diagram | Not reported (no figures) |
| 14a | Descriptive data: give characteristics of study participants (e.g., demographic, clinical, social) and information on exposures and potential confounders | Results, Sample characteristics (age, education, employment, sex and county by insurance status, with unadjusted tests) |
| 14b | Indicate number of participants with missing data for each variable of interest | Partially reported: Methods, Quantitative analysis (item-level missingness below 2%). The number of participants with missing data for each variable is not given |
| 15 | Outcome data: report numbers of outcome events or summary measures | Results, Sample characteristics (enrollment status counts); Primary barriers to enrollment among the uninsured and Table 1 (barrier counts and percentages with 95% CIs) |
| 16a | Main results: give unadjusted estimates and, if applicable, confounder-adjusted estimates and their precision (e.g., 95% confidence interval). Make clear which confounders were adjusted for and why they were included | Partially reported: Results, Socio-demographic predictors of enrollment and Table 3 (adjusted ORs with 95% CIs and p values); Results, Sample characteristics (unadjusted comparisons by insurance status); covariates listed in Methods, Quantitative analysis. Unadjusted odds ratios and the rationale for covariate selection are not reported |
| 16b | Report category boundaries when continuous variables were categorized | Methods, Quantitative analysis; Table 3 (education and employment categories; age analyzed as continuous) |
| 16c | If relevant, consider translating estimates of relative risk into absolute risk for a meaningful time period | Not reported |
| 17 | Other analyses: report other analyses done (e.g., analyses of subgroups and interactions, and sensitivity analyses) | Results, Primary barriers to enrollment among the uninsured and Table 1 (county comparison: chi-squared test and Cramér's V) |
| **Discussion** |  |  |
| 18 | Key results: summarize key results with reference to study objectives | Discussion, Principal findings |
| 19 | Limitations: discuss limitations of the study, taking into account sources of potential bias or imprecision. Discuss both direction and magnitude of any potential bias | Discussion, Strengths and limitations (including the likely underestimation of distrust as a contributing barrier, the small margin between distrust and cost, the likely direction of recruitment and household selection biases and mixed-mode effects). Analyses were not adjusted for clustering by community (Discussion, Strengths and limitations). The magnitude of potential biases is not quantified |
| 20 | Interpretation: give a cautious overall interpretation of results considering objectives, limitations, multiplicity of analyses, results from similar studies, and other relevant evidence | Discussion, Trust as an enabling resource: theoretical implications; Comparison with prior literature; Policy implications; Conclusions |
| 21 | Generalizability: discuss the generalizability (external validity) of the study results | Discussion, Strengths and limitations (four-county coverage; limited generalizability to non-fragile settings); Methods, Quantitative strand: sampling and survey (sample not nationally representative) |
| **Other information** |  |  |
| 22 | Funding: give the source of funding and the role of the funders for the present study and, if applicable, for the original study on which the present article is based | Declarations, Funding (no specific grant received) |
| **No.** | **Item** | **Where reported in this manuscript** |
| 1 | Describe the justification for using a mixed methods approach to the research question | Methods, Study design (distinct roles of the quantitative strand for prevalence, rank order and predictors, and the qualitative strand for meaning and subtypes of distrust); Introduction |
| 2 | Describe the design in terms of the purpose, priority and sequence of methods | Methods, Study design (concurrent design; strands conducted in parallel with equal priority; integration at interpretation) |
| 3 | Describe each method in terms of sampling, data collection and analysis | Methods, Quantitative strand: sampling and survey (four-stage stratified household survey, structured questionnaire); Qualitative strand: sampling and data collection (key informant interviews with a semi-structured guide and focus group discussions, audio-recorded, transcribed and de-identified); Quantitative analysis (double entry, complete-case analysis); Qualitative analysis. Further qualitative detail in Supplementary File 1 (COREQ) |
| 4 | Describe where integration has occurred, how it has occurred and who has participated in it | Partially reported: Methods, Study design and Integration (at interpretation stage, through a joint display and meta-inferences). Who participated in integration is not stated |
| 5 | Describe any insight gained from mixing or integrating methods | Results, Subtypes of institutional distrust: qualitative findings and Table 2 (joint display); Socio-demographic predictors of enrollment (final paragraph); Locating institutional trust in the Andersen framework and Table 4; Discussion, Principal findings |
| 6 | Describe any limitation of one method associated with the presence of the other method | Partially reported: Methods, Quantitative strand: sampling and survey and Discussion, Strengths and limitations (trust subtypes derived only from the qualitative strand, so no quantitative subtype estimates and no trust variable in the regression). Other limitations arising from combining the methods are not discussed |

Item wording follows the STROBE statement for cross-sectional studies (von Elm et al. 2007) and the GRAMMS guideline (O'Cathain et al. 2008). Locations refer to section headings and tables in the main manuscript. "Partially reported" indicates that the item is addressed in part, with the missing element stated; "Not reported" indicates that the item is not described in the manuscript.

